# Positive Autism Screening in Children Born Preterm

**DOI:** 10.1101/2025.04.04.25325278

**Authors:** Nuria Lisset Ontiveros Perez, Pamela Rios, Stella Firth Wang, Virginia A. Marchman, Ramkumar Aishworiya, Heidi M. Feldman

## Abstract

**Purpose:** Autism is more common among children born preterm than term. This study determined the prevalence of positive autism screening among 18-30 month old preterm-born children and evaluated sociodemographic, clinical, and neurodevelopmental factors associated with positive screens.

**Methods:** Secondary analyses of Stanford data from California Perinatal Quality Care Collaborative. Infants born < 32 weeks gestation between 2016-2020, who attended High-Risk Infant Follow-Up at 18-30 months, were classified into two groups based on results of Modified Checklist for Autism in Toddlers-Revised with Follow-Up (M-CHAT-R/F): positive-screen (score > 2) and negative-screen (≤ 2). We compared sociodemographics, clinical factors, and language development across groups.

**Results:** Prevalence of positive-screens was 12.2% (41/337). Children in the positive-screen group had lower gestational age, birthweight and longer hospital stays than children in the negative-screen group (all *p* < .05); gestational age was the primary factor associated with a positive screen (OR 0.75, 0.56-0.99). We found no group differences in sociodemographics or medical complications; children in the positive-screen group had lower language scores than children in the negative-screen group (*p* < .001). All children in the positive-screen group and ~⅓ of children in the negative-screen group scored low in language.

**Conclusion:** The high prevalence of positive-screens reinforces the importance of early screening for autism in very preterm children. Gestational age at birth was the only factor associated with positive-screens. Language difficulties were not specific to children with positive-screens, highlighting the need for autism screening and routine developmental assessments for preterm children.

---

Positive Autism Screening in Children Born Preterm Prematurity, defined as birth prior to 37 weeks of gestation, is a highly prevalent condition, affecting about 10% of live births. Prematurity is associated with many neurodevelopmental and neurobehavioral conditions, including autism. Estimates of autism prevalence in the general US population ranged from 1.1% to 2.8% from 2008 to the present (CDC, 2020; Baio, 2019). Autism prevalence in children born preterm ranged from 2.6% - 18.5% during a comparable time frame (Marín Soro et al., 2024; Crump et al., 2021; Gray et al., 2015; Kuzniewicz et al., 2014; Moore et al., 2012). In studies that directly compared pre- and full-term children, the prevalence of autism was consistently higher in preterm samples (Crump et al., 2021; Gray et al., 2015). Autism screening is typically a routine component of protocols designed to monitor neurodevelopment in children born preterm after their hospital discharge.

The extant medical literature has not clearly identified the specific reasons for this higher incidence of autism among preterm children. Factors associated with autism in children born preterm have included gestation age at birth, sex, and sociodemographic variables (Gray et al., 2015, Kuzniewicz et al., 2014, Moore et al., 2012) but vary across studies. Identification of specific medical factors associated with the eventual development of autism could potentially facilitate targeted identification of such children at a younger age and early initiation of appropriate treatments. Along this line, better delineation of the clinical profile, including developmental status of preterm children who are more likely to have autism could enable the provision of timely intervention and parental counselling. At present, other developmental markers, including language deficits in these children, and how these are associated with autism screening results are not well described.

In California, the High Risk Infant Follow-Up (HRIF) clinics, administered through <u>California Children’s Services (CCS),</u> provide outpatient longitudinal developmental care, including autism screening, to infants born before 32 weeks of gestation or with birth weight less than 1500 grams (very preterm birth). Standardized data forms, summarizing the results of a comprehensive assessment, are sent from individual clinics to California Perinatal Quality Care Consortium (CPQCC), creating detailed records for each child at each site that link sociodemographic characteristics, clinical data, and medical complications to the results of developmental testing and autism screening. At the Stanford High Risk Infant Follow-Up clinic, the Modified Checklist for Autism in Toddlers-Revised with Follow-Up (M-CHAT-R/F) is typically administered between 18-30 months of age with the recommended interview (Robins et al., 2014). At the same visit, children are also assessed for language development.

A review of the CPQCC records of children attending the Stanford High Risk Infant Follow-Up (HRIF) clinic thus afforded us an opportunity to evaluate results of autism screening in a sample of children born preterm. We asked the following research questions:

- What proportion of children born very preterm had positive results on autism screening between 18-30 months of age?
- What specific sociodemographic (e.g., sex, health insurance) and clinical factors (e.g., gestational age, birth weight, and presence of specific medical complications) were associated with a positive screen of autism?
- How did the scores on language assessment differ between children who had positive versus negative autism screening results?

Analyzing factors associated with an elevated likelihood of autism, as indicated by positive screening results, may provide insights into the reasons for the high prevalence in this population and may contribute to refining screening protocols for children born preterm. We hypothesized that child sex and clinical factors, including the presence of medical complications of prematurity, would be associated with positive autism screening results. We also hypothesized that presence of language delay would be associated with positive screening results.

## Methods

### Design and Participants

Secondary data analysis of children followed at the Stanford Medicine Children’s Health HRIF clinic included in the CPQCC dataset. Inclusion criteria were very preterm birth (< 32 weeks gestational age at birth or birth weight < 1500 grams); birthdate between 2016-2020; age of assessment between ages 18-30 months, adjusted for the degree of prematurity (assessments between 2017-2023); and results for the M-CHAT-R/F recorded at that visit. Exclusion criteria were any children with congenital malformations or disorders, blindness, or deafness.

### Procedure

At the Stanford HRIF clinic, the Modified Checklist for Autism in Toddlers-Revised with Follow-Up (M-CHAT-R/F) is typically administered between 18-30 months of age, with the recommended interview conducted if needed, for respondents with initial scores between 3-7 (Robins et al., 2014). We classified the participants in two groups based on scores on the M-CHAT-R/F: (1) positive-screens, indicating an elevated likelihood of autism, defined as scores > `2 after interview and (2) negative-screens, indicating low likelihood of autism defined as initial scores ≤ 2 or post-interview scores < 2`. We extracted the following socioeconomic data from each child’s record: sex, child age at assessment, mother’s age, singleton vs multiple, race, ethnicity, caregiver education, language spoken at home, and insurance. We examined the following general clinical factors: gestational age at birth, birth weight, and length of neonatal hospital stay. We extracted data regarding three prevalent medical conditions associated with prematurity that indicated significant medical complications and might therefore have increased the likelihood of autism: neurological abnormalities, retinopathy of prematurity, and persistent chronic lung disease. We classified a child as having neurological complications if their record indicated a positive on one or more of three variables included in the dataset: “any neurological abnormality”, “intracranial hemorrhage”, or “intracranial pathology with potential for adverse neurologic outcome”. We determined that the child had retinopathy of prematurity if there was a positive response for “the presence of a history of retinopathy” without or with visual impairment. Finally, we determined the presence of chronic lung disease based on whether they had a positive response to one or more of four conditions: (1) “inhaled daily bronchodilators”, (2) “inhaled intermit use of bronchodilators”, (3) “were on oxygen”, or (4) “oxygen was required for more than 28 days of life with chronic lung disease”.

We also extracted scores from the Clinical Linguistic and Auditory Milestone Scale (CLAMS) of the Capute Scales of Infant Development (Accardo, 2005). Language assessment is conducted at the same visit as the M-CHAT-R/F. We divided the sample into “high language,” defined as developmental quotient (DQ) score of > 85, i.e., within one SD of the mean, and “low language,” defined as a DQ score of ≤ 85, i.e., greater than one standard deviation below the mean. Supplementary Table 1 includes the exact variable names for all data analyzed. Data Analysis

To derive descriptive statistics, demographic factors, clinical factors and medical complications, as well as language development scores were compared as a function of screening status. For statistical analysis, we compared screening groups on these variables using independent sample t-tests and chi-square tests, as appropriate. We considered multiple factors related to a positive screening result using logistic regression to derive odds ratios. All analyses were run in R (Version 4.4.1).

## Results

Figure 1 presents the consort diagram. Of the 553 children born very preterm and registered as part of HRIF from Stanford, 88.4% were seen at 18-30 months of age. Of those, 337 or 68.9% had results of the M-CHAT-R/F in their record. The mean age for M-CHAT-R/F administration and language assessment was 24.5 months, adjusted for the degree of prematurity. A total of 41 of the 337 (12.2%) had positive-screens and 89.8% had negative-screens.

**Figure 1.**
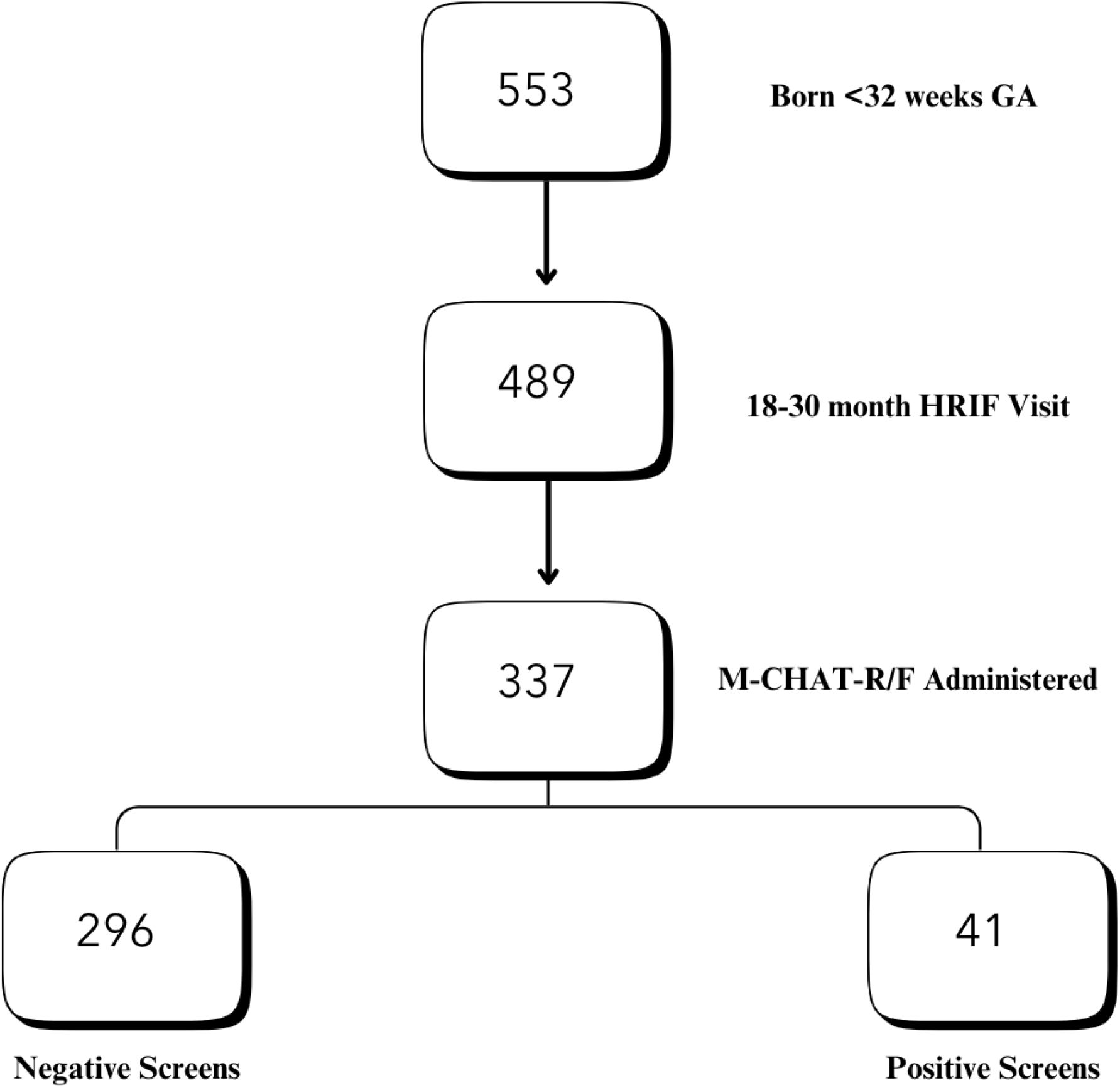
Consort Diagram representing the sample from the Stanford High Risk Infant Follow-Up (HRIF) dataset of children born 2016-2020.

Table 1 presents descriptive and comparative data for the sociodemographic factors as a function of screening results group. There were no statistically significant differences in incidence by sex, child age, singleton birth, race, ethnicity, caregiver education, language spoken at home, or use of public insurance between the 2 groups (all *p* > .20). Caregiver age was statistically higher in the positive-screens vs negative screens group (M = 33.3, SD = 5.2 vs M = 31.6, SD = 6.1, *p* = 0.05), though the mean age for both groups was over 31 years.

**Table 1.** Demographic, clinical factors and medical complications associated with M-CHAT-R/F negative and positive screens.

| Variables | M-CHAT-R/F<br>Negative Screens<br>% or M(SD) | M-CHAT-R/F<br>Positive Screens<br>% or M(SD) | $\chi^2$ or $t$ | p |
| --- | --- | --- | --- | --- |
| **Demographics** |  |  |  |  |
| Child Sex (Male) | 58% | 66% | 1.16 | 0.28 |
| Child's Age at Assessment (Months) | 24.5 | 24.5 | -0.06 | 0.94 |
| Mother's Age (Years) | 31.6(6.07) | 33.3(5.21) | -1.98 | 0.05 |
| Singleton and Multiple births (Singleton) | 75% | 76% | 9.11 | 1.00 |
| Race (Non-white) | 61% | 59% | 0.26 | 0.61 |
| Ethnicity (Hispanic) | 47% | 51% | 0.49 | 0.48 |
| Caregiver Education (≤ Highschool) | 26% | 29% | 0.92 | 0.33 |
| Language Spoken at Home (Non-English) | 29% | 32% | 0.70 | 0.40 |
| Insurance (Public) | 61% | 68% | 2.08 | 0.35 |
| **Clinical Factors** |  |  |  |  |
| Gestational Age at birth | 28.3(2.31) | 27.1(2.49) | 3.04 | <.01 |
| Length of Stay | 71.7(45.4) | 97.2(63.4) | -2.52 | 0.01 |
| Birth Weight | 1193.1(380) | 1067.8(371) | 2.04 | 0.05 |
| **Medical Complications** |  |  |  |  |
| Neurologic Abnormalities | 21% | 24% | 0.03 | 0.86 |
| Retinopathy of Prematurity | 64% | 71% | <.01 | 0.96 |
| Chronic Lung Disease | 36% | 37% | 1.75 | 1.00 |

Table 1 also presents descriptive data for three global clinical factors: gestational age, birth weight, and length of hospital stay. Children in the positive-screen group had lower gestational age (M = 27.1 (2.5) vs M = 28.4 (2.3)) and birthweight (1068g (371) vs 1193 (380)) and longer hospital stays (97.2 (63.4) vs 72 days (45.4)) (all *p* < .05) than children in the negative-screen group. In a logistic regression analysis, predicting screening result group from all three general clinical variables simultaneously, gestational age was the only significant factor (*p* = 0.04). Birth weight and length of stay were no longer significant when gestational age was included in the model (*p* = 0.22). The odds ratio (OR) for gestational age was 0.75 (95% CI [0.56, 0.99]) indicating that the odds of a positive screen for autism decreased by 25% for every additional week of gestation. The OR for birth weight was 1.01 (95% CI [0.99, 1.03]) and for length of stay was 1.04 (95% CI [0.99, 1.01]), indicating that these factors did not significantly change the odds of autism risk after consideration of gestation age.

None of the three specific medical conditions associated with prematurity were associated with the screening group. Among children with neurological complications (n = 73), 10 (13.7%) were classified as positive screens. However, children with a positive screen were not significantly more likely to have neurological complications than children with negative screens. Among children with retinopathy of prematurity (n = 29), 4 (13.7%) were classified as positive screen but retinopathy was equally frequent in both screen groups. Among children with chronic lung disease (n = 123), 15 (12.2%) were classified as positive screens, but chronic lung disease was not associated with an elevated likelihood of autism.

A total of 117 children were assessed using the CLAMS. The positive-screens group had lower mean DQ language scores (M = 53.9 (18.1)) than the negative screen group (M = 92.5 (20.1), t = 7.81, df = 21.28, p < .001). Figure 2 shows the number of children with low language (≤ 85) and high language (> 85) as a function of autism screening results. Children in the negative-screen group fell into both low language (n = 37, M = 71.4 (9.40)) and high language groups (n = 64, M = 105.0 (13.2)). No child in the positive-screen group had language scores > 85, i.e., one standard deviation above the mean (*X*^2^ = 96.1, df = 65, p < .01).

**Figure 2.**
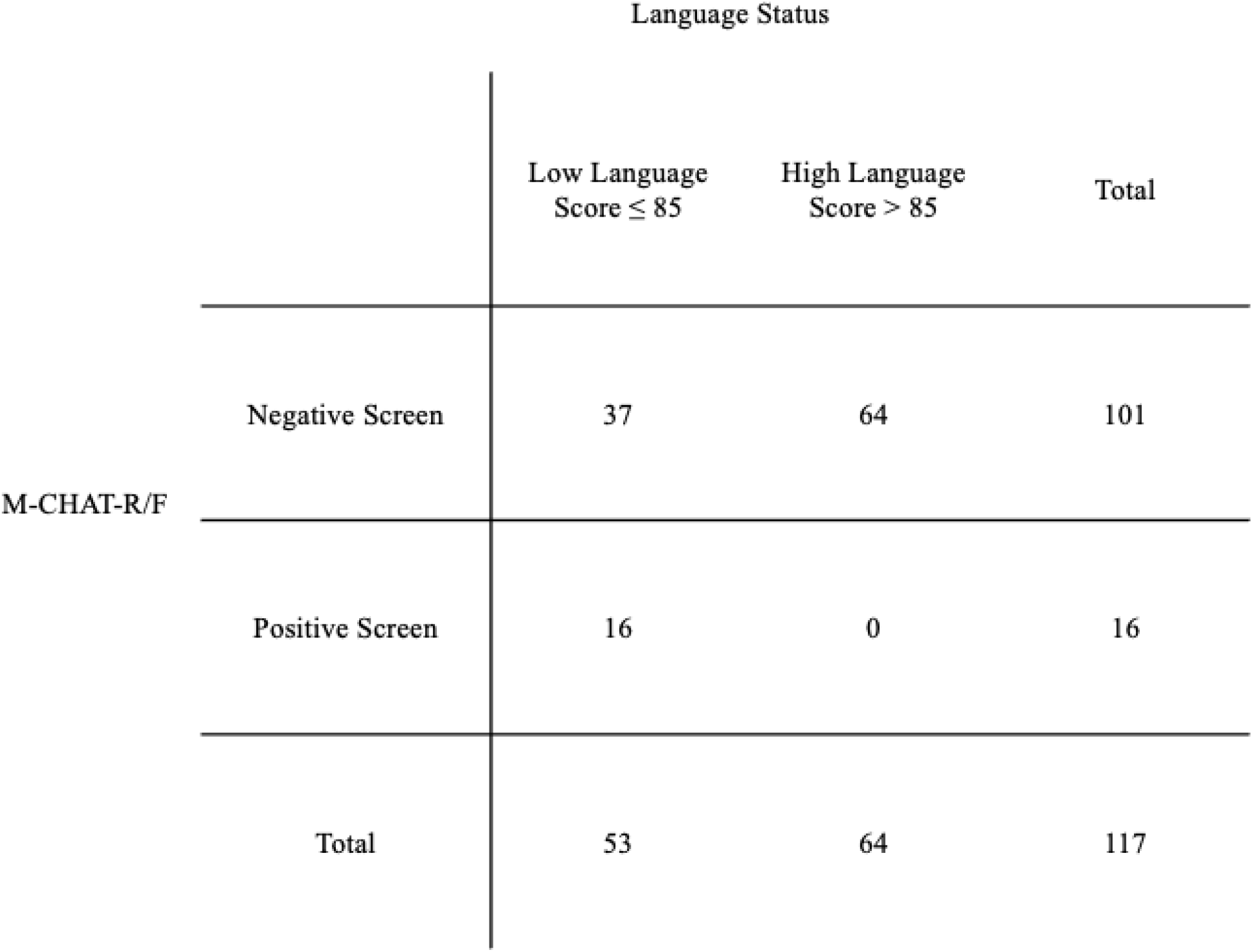
Capute language scores in relation to the results on the M-CHAT-R/F screening for autism in children born very preterm.

## Discussion

The overall prevalence of positive screening for autism in this sample of very preterm children was nearly 3 times higher than that reported for children in the general population (Robins et al., 2014). We recognize that many of these children may not have been diagnosed with autism after a comprehensive assessment. We do not have information on which children were diagnosed with autism in this data set. However, the results suggest that at least 12% of very preterm children need further definitive evaluation for autism. The precise proportion of high likelihood of autism found here may be inflated compared to other studies because the threshold for positive screens included moderate and high risk of autism (Robins, 2014). Nonetheless, programs that monitor development in children born preterm must be prepared to conduct autism evaluations on a routine basis. Further, given that the positive predictive value of the M-CHAT-R/F is high, at 90% for any developmental condition (such as language delay), these data also suggest that at least 12% of a preterm sample will likely warrant specific developmental interventions and follow-up (Robins et al., 2014; Aishworiya et al., 2023).

Contrary to our hypothesis, we found no association between sociodemographic variables and the likelihood of screening positive for possible presence of autism. Within the general population, the prevalence of autism among boys is generally 3 to 4 times the prevalence among girls (Moore et al., 2012). In this study, the sex differences were no longer present. Associations of social class and autism have varied across studies (Gray et al., 2015). We did not find an association with our measure of socioeconomic status.

As hypothesized, we found significant associations of positive-screen results with gestational age at birth, birth weight, and length of neonatal hospital stay. However, only gestational age remained significant when all three variables were considered simultaneously as predictors. The odds ratio analysis indicated that the risk of a positive screen for autism dropped by 25% for every additional week of gestation. Contrary to the hypotheses, none of the specific common medical complications of preterm birth—neurological complications, retinopathy of prematurity, and chronic lung disease–were associated with positive-screens. These findings suggest that general factors associated with preterm birth, such as prolonged hospitalization, sensory abnormalities in neonatal intensive care units, repeated painful procedures, and lack of a consistent family presence may be more important in the development of autism than any specific health condition or than sex and socioeconomic status.

Low scores on language testing were associated with positive screens for autism. These findings are not surprising because early developmental indicators of autism include delays in language development. Moreover, language, cognitive, and motor scores are highly correlated among children born preterm (Loeb et al., 2020). However, approximately half of the children with low language scores had negative screens for autism. This finding suggests that low language scores by themselves do not necessarily imply the presence of autism; instead, preterm infants may have low language scores without the presence of behaviors consistent with autism. Thus, when designing clinical care pathways for follow-up of preterm children, testing both language status and autism likelihood are appropriate for this population.

This study has limitations. First, the data are derived from a single center. Future studies could expand these analyses to multiple sites within this state-wide data set of children born preterm. Secondly, the dataset had missing data due to the fact that the data collection period included the COVID pandemic. During that period, many planned HRIF visits were postponed or cancelled. Many visits were conducted via telehealth and a M-CHAT-R/F paper screening form, used at that time, was not routinely collected. Also, early in the pandemic, clinicians did not attempt developmental testing, leading to considerable missing results for language development. Analyses should be replicated in an era in which in-person visits allow parental completion of the M-CHAT-R/F and reliable developmental testing for the entire study cohort.

## Conclusion

Our study illustrates a high incidence of positive-screening results for autism in a sample of children born very preterm, with gestational age at birth being the primary factor associated with this elevated likelihood of autism. These data suggest that general factors associated with preterm birth rather than any specific medical complication or sociodemographic factor may be associated with a developmental trajectory toward autism. Further, our results suggest that language delays and behavioral characteristics of autism are not tightly linked, given that about half of the sample with low language scores did not have positive screening results for autism. Though results should be replicated, they reinforce the importance of early screening for autism in all very preterm children.

## Data Availability

All data produced in the present study are available upon reasonable request to the authors.

https://osf.io/2k57e

## Statements and Declarations

### Declarations

All authors contributed to the study conception and design. Material preparation, data collection and analysis were performed by Nuria Lisset Ontiveros Perez, Pamela Rios, and Virginia Marchman. The first draft of the manuscript was written by Nuria Lisset Ontiveros Perez and all authors commented on previous versions of the manuscript. All authors read and approved the final manuscript.

## Acknowledgements

This research work was supported by grants from the National Institutes of Health-Eunice Kennedy Shriver National Institute of Child Health and Human Development (HM Feldman, PI; 2RO1- HD069150) and National Medical Research Council, Singapore (R Aishworiya, PI Transition Award (MOH-001579)).

## Compliance with Ethical Standards

### Ethics approval

This retrospective chart review study involving human participants was in accordance with the ethical standards of the institutional and national research committee and with the 1964 Helsinki Declaration and its later amendments or comparable ethical standards. The Human Investigation Committee (IRB) of Stanford University approved this study.

### Consent

The IRB determined that formal consent was not necessary for this retrospective, low risk study.

---

Supplementary Table 1. CPQCC Variable Names Used in the Analysis, available at: https://osf.io/2k57e

